# Integrative Multi-Omics Framework for Causal Gene Discovery in Long COVID

**DOI:** 10.1101/2025.02.09.25321751

**Authors:** Sindy Pinero, Xiaomei Li, Lin Liu, Jiuyong Li, Sang Hong Lee, Marnie Winter, Thin Nguyen, Junpeng Zhang, Thuc Duy Le

## Abstract

**Background:** Long COVID, or Post-Acute Sequelae of COVID-19 (PASC), involves persistent, multisystemic symptoms in about 10–20% of COVID-19 patients. Although age, sex, ethnicity, and comorbidities are recognized as risk factors, identifying genetic contributors is essential for developing targeted therapies.

**Methods:** We developed a multi-omics framework using Transcriptome-Wide Mendelian Randomization (TWMR) and Control Theory (CT). This approach integrates Expression Quantitative Trait Loci (eQTL), Genome-Wide Association Studies (GWAS), RNA sequencing (RNA-seq), and Protein-Protein Interaction (PPI) networks to detect causal genes and regulatory nodes that drive critical expression changes in Long COVID.

**Results:** We identified 32 causal genes (19 previously reported and 13 novel), which act as regulatory drivers influencing disease risk, progression, and stability. Enrichment analyses highlighted pathways linked to the SARS-CoV-2 response, viral carcinogenesis, cell cycle regulation, and immune function. Analysis of other pathophysiological conditions revealed shared genetic factors across syndromic, metabolic, autoimmune, and connective tissue disorders. Using these genes, we identified three distinct symptom-based subtypes of Long COVID, offering insights for more precise diagnosis and potential therapeutic interventions. Additionally, we provided an open-source Shiny application to enable further data exploration.

**Conclusion:** Integrating TWMR and CT revealed genetic mechanisms and therapeutic targets for Long COVID, with novel genes informing pathogenesis and precision medicine strategies.

## 1 Introduction

Long COVID, also known as Post-Acute Sequelae of COVID-19 (PASC), is a complex condition marked by the persistence or development of symptoms as a result of SARS-CoV-2 infection. Long COVID is defined differently by various organizations. For instance, the World Health Organization (WHO) and the Centers for Disease Control and Prevention (CDC) describe it as symptoms persisting three months after infection and lasting for at least two months [1, 2], whereas the National Institute for Health and Care Excellence (NICE) considers it to begin as early as one-month post-infection [3–5]. Regardless of the definition timing, key risk factors include age, sex, ethnicity, socioeconomic status, vaccination status, smoking, and underlying health conditions [6]. Moreover, studies have linked various biomarkers to Long COVID, particularly those related to inflammation, immune dysfunction, and coagulation abnormalities [7].

Despite significant progress in identifying risk factors and clinical markers [6, 7], understanding the role of gene expression as a causal factor in Long COVID remains a major challenge. This gap in knowledge presents a significant barrier to developing and implementing interventions and targeted therapies [8], highlighting the need for novel approaches focusing on gene expression patterns that may contribute to Long COVID. Identifying these Long COVID-causing genes is essential for advancing targeted treatment strategies. It could also improve diagnostic accuracy and promote better monitoring and prediction of patient outcomes [9].

Computational methods for identifying disease-causing genes typically follow two primary strategies, each offering distinct advantages that complement the other. The first strategy aims to identify genes associated with disease risk and prevention, often using approaches such as Transcriptome-Wide Mendelian Randomization (TWMR) [10]. The TWMR approach incorporates transcriptomic data into MR studies by using genetic variants that affect gene expression, such as Expression Quantitative Trait Loci (eQTLs), to establish causal relationships between gene activities and diseases. The TWMR methodology can identify whether altered gene expression directly influences an outcome—in this instance, Long COVID—and reveal potential therapeutic targets. The resulting analysis reveals which genetic factors influence disease susceptibility or protection through genetic associations and causal inference, allowing researchers to identify specific genetic variants with direct causal effects on diseases. However, TWMR analysis often requires strong genetic instruments (e.g., Single Nucleotide Polymorphisms (SNPs) that robustly modulate gene expression), and determining causal relationships becomes more complex when facing confounding variables or pleiotropy.

The second strategy identifies genes or proteins that are crucial in biological networks. Techniques such as Bayesian Networks [11], Node Importance [12], and the Control Theory (CT) [13] are used to understand how different genes and proteins interact within biological pathways, considering the interconnected nature of biological systems. CT is particularly useful for identifying critical nodes or key genes and proteins significantly influencing the entire network. By finding these critical nodes (network driver genes), researchers can determine which components are most effective to target to stabilize or control disease-related disruptions. For example, CT methods have been used in cancer research to identify key regulatory genes like *TP53* whose modulation can restore network stability, thereby providing focused therapeutic opportunities [14].

In this study, we propose a novel framework to explore and discover potential genes involved in Long COVID by integrating two complementary strategies: MR [10] and CT [13], along with multi-omics data, including eQTLs, Genome-Wide Association Studies (GWAS), RNA sequencing (RNA-seq), and human Protein-Protein Interaction (PPI) network. Our framework investigates candidate causal genes that may contribute to Long COVID risk and examines their potential regulatory roles within a network. Specifically, we discover genes whose expression patterns suggest either increased susceptibility to Long COVID or a crucial role in maintaining biological network stability. By integrating these methodologies and utilizing multi-omics data, our analysis provides comprehensive insights into the potential genetic mechanisms underlying Long COVID and highlights candidate therapeutic targets for further investigation.

Our study has identified 32 causal genes for Long COVID, of which 19 have been confirmed by existing literature, supporting the effectiveness of our findings. The remaining genes represent promising candidates for follow-up experiments. Among these candidates, we discovered driver genes that act as risk or preventive factors and network driver genes that regulate and stabilize disease network structure. Enrichment analyses revealed important biological pathways in Long COVID, including SARS-CoV-2 infection, viral carcinogenesis, cell cycle regulation, and immune response mechanisms. Using the identified causal genes, we clustered Long COVID patients into three distinct subtypes with different symptom profiles, establishing a foundation for personalized diagnostic and therapeutic approaches. This work represents a significant step toward customized management and treatment strategies for Long COVID, ultimately improving patient outcomes.

To facilitate the application of our framework, we developed a web application (Shiny app) that allows users to generate gene lists by adjusting parameters related to direct (MR) and network-based (CT) causal approaches. This tool provides researchers and clinicians with an accessible platform to explore parameter variations and analyze their data, enhancing the reproducibility of our findings.

## 2 Results

### 2.1 Overview of the Causal Gene Discovery Framework

The causal gene discovery framework integrates diverse data sources, including eQTL, GWAS, RNA-seq, and PPI networks, to identify genes with causal roles in Long COVID (Fig. 1). It begins by processing multi-omics input data (Fig. 1A) and then applies an integrative scoring method (Fig. 1B) that combines TWMR with network analysis using CT. This approach balances the contributions of risk/preventive factors and network-critical genes through a parameter (*α*) that can be adjusted depending on the goals. The output (Fig. 1C) ranks causal genes based on the weighted scores, offering insights into their roles within the Long COVID network. Finally, downstream analyses (Fig. 1D), including Enrichment Analysis (EA), literature validation, and subtype identification, help discover disease mechanisms and prioritize therapeutic targets. This comprehensive computational approach integrates genetic and network-based perspectives, providing deeper insights into the nature of Long COVID.

**Fig. 1:**
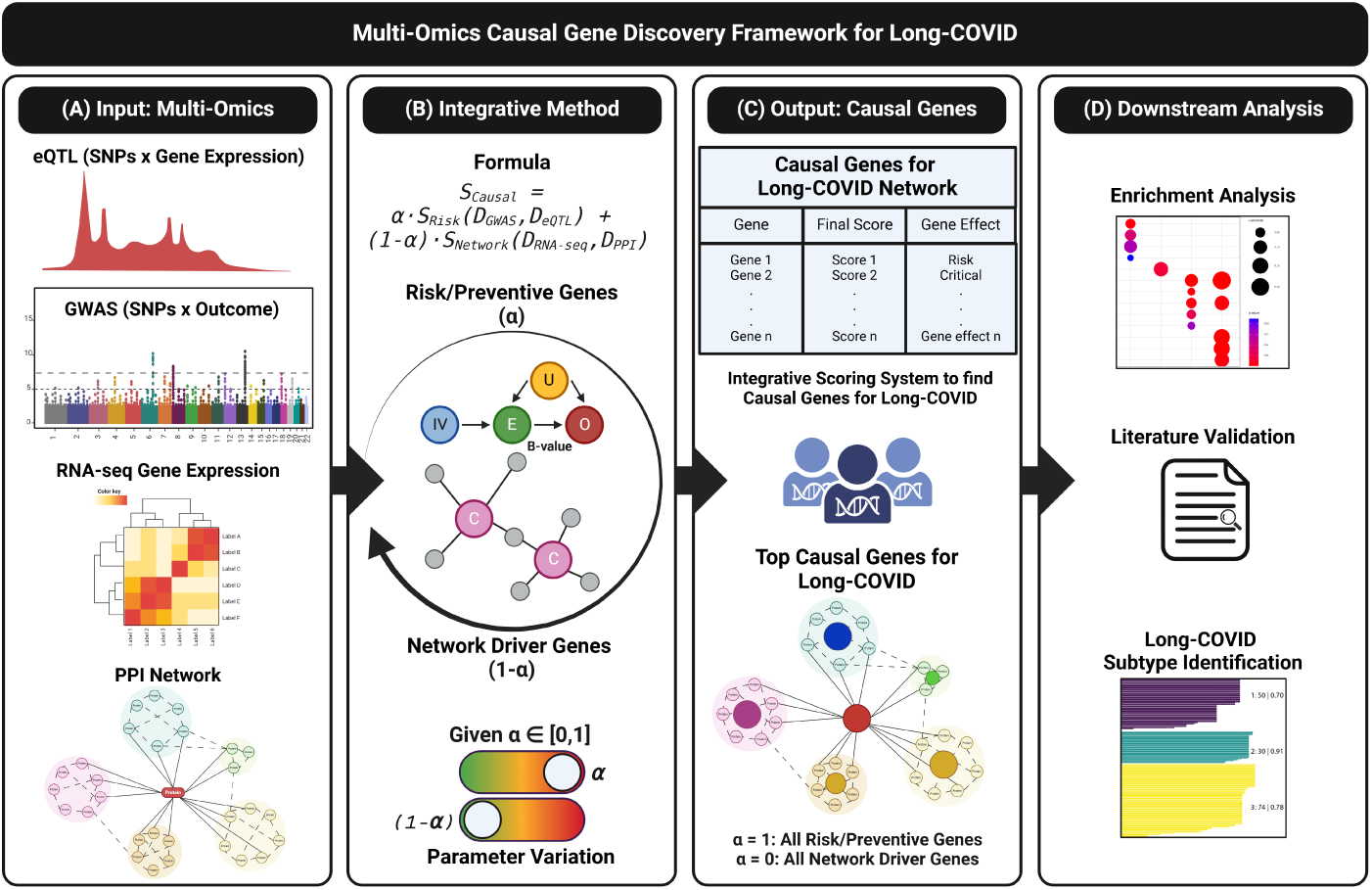
A causal gene discovery framework for Long COVID using multi-omics data. (A) The input data includes expression Quantitative Trait Loci (eQTL), Long COVID Genome-Wide Association Studies (GWAS), RNA sequencing (RNA-seq), and the human Protein-Protein Interaction (PPI) network. (B) A fusion approach to evaluating gene expression by integrating Transcriptome-Wide Mendelian Randomization (TWMR) and Control Theory (CT) scores. (C) Significant genes are ranked based on the weighted scores. (D) Downstream analyses include Enrichment Analysis (EA), literature review, and Long COVID subtype identification. SNPs: Single Nucleotide Polymorphisms, IVs: Instrumental Variables. E: Exposure. O: Outcome. U: Confounders.

By treating genetic variants as instrumental variables (IVs), two-sample MR methods detect genetically regulated risk exposures for complex diseases using only summary statistics. When considering gene expression as an exposure in TWMR analyses, we aim to identify gene expressions that have causal relationships with the disease of interest. In our case, we focus on identifying the genes that act as risk or protective factors for Long COVID. Given the limited number of eQTLs available as IVs for a gene, which makes detecting invalid IVs challenging, we adopt the multi-tissue approach, *Mt-Robin* [10]. This method uses eQTL data in a mixed model to identify IV-specific random effects due to pleiotropy arising from estimation errors in eQTL summary statistics, enabling accurate inference of the dependence (fixed effects) between eQTL and GWAS effects, even in the presence of invalid IVs.

While MR approaches identify genes that directly affect Long COVID, network biology approaches, such as CT, have shown that genes driving the disease are not limited to those directly linked to disease phenotypes [13]. In this work, we employ a CT approach to extract a list of genes that serve as network drivers for Long COVID—i.e., genes whose removal or intervention would disrupt the biological networks associated with the disease, thereby affecting disease outcomes. These network driver genes may or may not have direct causal relationships with the disease.

To create a comprehensive list of causal genes for Long COVID and understand their roles in regulating the disease, we use a fusion approach that integrates the two methods described above (see Methods for details). Specifically, we calculate the scores of each gene using the following formula:

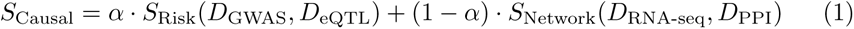

where:

- *S*_Causal_ represents the final score of each gene.
- *S*_Risk_(*D*_GWAS_*, D*_eQTL_) is the score derived from the TWMR approach (*Mt-Robin*) to identify risk and protective causal genes using GWAS and eQTL datasets.
- *S*_Network_(*D*_RNA-seq_*, D*_PPI_) is the score from the CT approach to identify network driver genes based on RNA-seq data and the human PPI network.
- The parameter *α* controls the contribution of each risk/protective causal gene, while 1 *− α* adjusts the influence of each network-critical gene.

The formula (1) integrates an approach that ranks genes by combining their causal effects and significance within the Long COVID network, providing a comprehensive prioritization based on both causal and network properties.

Thus, this causal multi-omics approach provides insights into Long COVID’s genetic mechanisms while highlighting possible intervention targets.

### 2.2 Dynamic Visualization of Long COVID Causal Genes: A Shiny Application

In our model, the parameter *α* serves as an adjustable coefficient that enables researchers to explore different scenarios for prioritizing protein-coding genes based on their roles in influencing disease risk or prevention and controlling the Long COVID network.

When *α* approaches 1, the model prioritizes genes linked to disease risk and preventive scenarios. These genes are directly associated with Long COVID pathogenesis, highlighting potential therapeutic targets for intervention.

Conversely, as *α* approaches 0, the model emphasizes driver genes critical to the disease network’s structure. These genes regulate key interactions within the network, positioning them as potential therapeutic targets to restore lost stability or modulate pathological states.

The model integrates both perspectives in the intermediate ranges of *α*, balancing the importance of network controllability with disease risk. In these cases, genes that significantly influence the network and are closely linked to disease risk become key players, making them important targets for further investigation.

Researchers can dynamically explore these shifts in gene rankings by adjusting *α* in our interactive tool available at Dynamic Causal Genes Visualization in a Long COVID Network^1^. This instrument allows a detailed examination of how genes transition from disease risk or preventive factors (*α →* 1) to network drivers (*α →* 0).

### 2.3 Causal Genes of Long COVID

By varying *α* values in our model, we identified a comprehensive set of causal genes for Long COVID with distinct roles. Fig. 2 shows the sets of these causal genes corresponding to specific values of *α*. As *α* approaches 1, the model outputs genes classified as risk (red) or preventive (green), inferred from the color coding of their effect sizes, with red representing positive effect sizes (risk) and green representing negative effect sizes (preventive). Decreasing *α* towards zero shifts the focus to network driver genes that control the Long COVID PPI network (yellow).

**Fig. 2:**
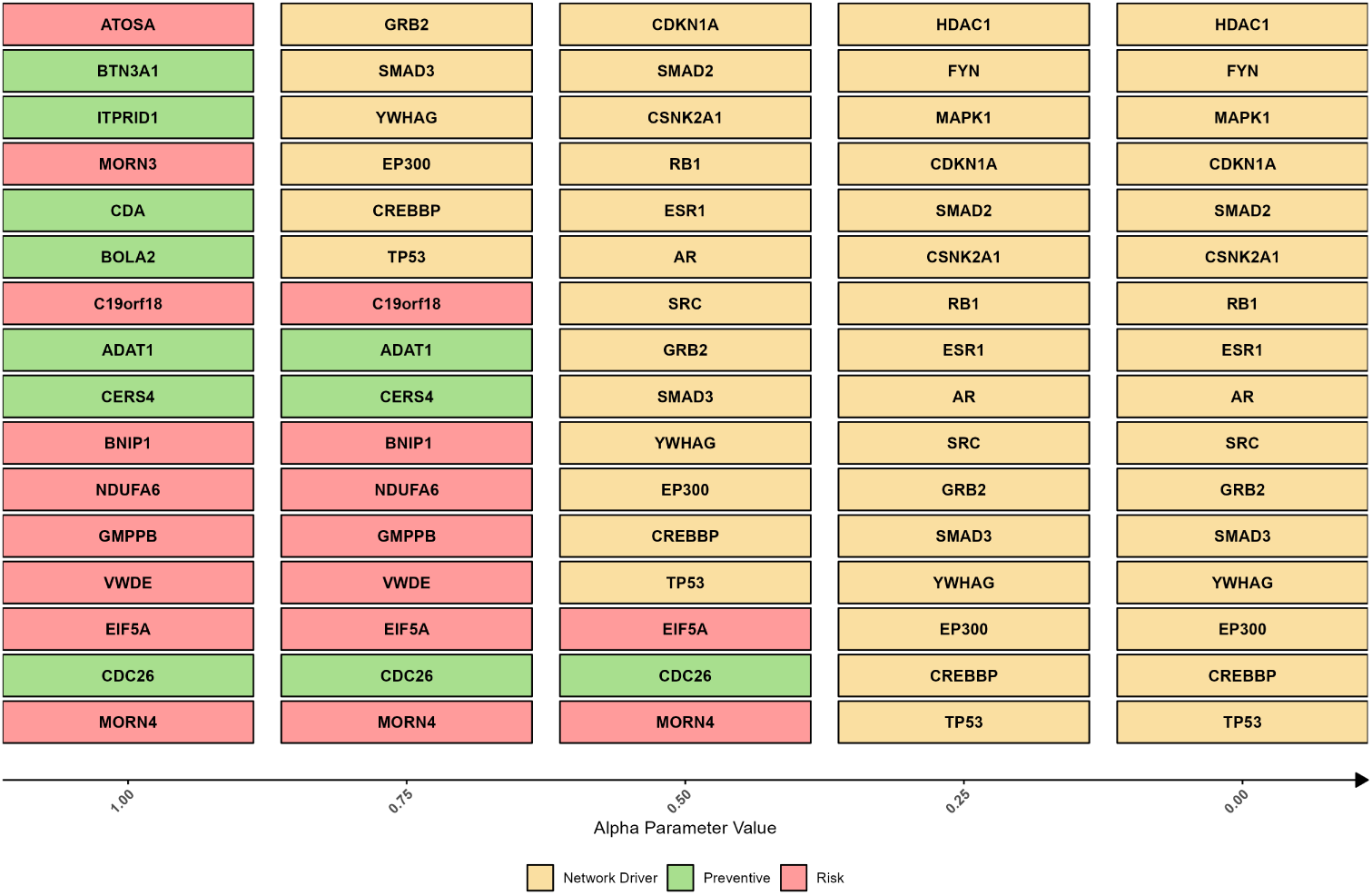
Top causal genes ranked by their final score. *S***_Causal_.** These genes, obtained from our framework, are sorted horizontally based on their absolute effect size in ascending order and classified vertically across different *α* values. The parameter *α* balances the direct effect of genes on the disease (*S*_Risk_) and their network controllability roles (*S*_Network_). At *α* = 1, the model outputs disease risk (red) and prevention (green) genes, while decreasing *α* towards 0, the focus shifts to network driver genes that control the biological network (yellow).

Genes like membrane occupation and recognition nexus repeat containing 4 (*MORN4* ), cell division cycle associated 26 (*CDC26* ), and eukaryotic translation initiation factor 5A (*EIF5A*) consistently rank highly across different *α* values (1.00 to 0.50), suggesting a strong causal relationship between their expression levels and disease risk or preventive mechanisms. Using SNPs as IVs in our analysis, we estimated the causal effects of the expression of these genes on Long COVID risk. The consistently high ranking of *MORN4*, *CDC26*, and *EIF5A* indicates that their expression levels may significantly contribute to disease susceptibility, making them potential key targets for intervention strategies focused on reducing disease risk (see Fig. 2). The complete list of SNPs used as IVs for each gene’s expression can be found in [Supplementary Data 1](https://github.com/SindyPin/Causal-Multiomics-Method/tree/main/SM/SM1).

As *α* decreases, the model shifts focus from the MR perspective to the CT perspective, prioritizing the balance between risk-related genetic contributions and network control dynamics. This transition highlights the framework’s flexibility in integrating these perspectives. Notably, genes such as tumor protein p53 (*TP53* ), cyclic adenosine monophosphate response element-binding protein-binding-protein (*CREBBP* ), early region 1A binding protein p300 (*EP300* ), tyrosine 3-monooxygenase/tryptophan 5-monooxygenase activation protein gamma (*YWHAG* ), SMAD family member 3 (*SMAD3* ), and the growth factor receptor-bound protein 2 (*GRB2* ) become increasingly crucial in the network, emphasizing their roles in maintaining network control (see Fig. 2, with these genes highlighted in yellow).

When considering the union of the top genes for each *α* value in our analysis, we identified 32 unique core causal genes for Long COVID. This comprehensive set of genes represents the most influential factors across the spectrum of our *α* parameter, which balances disease-related impact and network controllability.

Of these 32 genes, 19 have been previously identified in COVID-19 and/or Long COVID studies, reinforcing their importance in the disease process. These include well-known genes such as the androgen receptor (*AR*), butyrophilin subfamily 3 member A1 (*BTN3A1* ), cyclin-dependent kinase inhibitor 1A (*CDKN1A*), *CREBBP*, *EIF5A*, *EP300*, estrogen receptor 1 (*ESR1* ), atos homolog A (*ATOSA*), FYN proto-oncogene (*FYN* ), *GRB2*, histone deacetylase 1 (*HDAC1* ), mitogen-activated protein kinase 1 (*MAPK1* ), NADH:ubiquinone oxidoreductase subunit A6 (*NDUFA6* ), retinoblastoma transcriptional corepressor 1 (*RB1* ), SMAD family member 2 (*SMAD2* ), *SMAD3*, sarcoma proto-oncogene (*SRC* ), *TP53*, and *YWHAG*. These genes have been associated with various SARS-CoV-2 infection and Long COVID aspects, including roles as hub genes, drug targets, and factors influencing disease severity (Table 1). The high number of confirmed Long COVID genes suggests that our framework effectively identifies causal genes.

**Table 1:** Core causal genes for Long COVID confirmed by the literature. These 19 genes were validated by existing COVID-19 (COV) and/or Long COVID (LCV) studies, reinforcing our findings. Sev.: Severity, Reg.: Regulation, Polymo.: Polymorphisms. For more supporting literature, refer to [Supplementary Data 2](https://github.com/SindyPin/Causal-Multiomics-Method/tree/main/SM/SM2).

| Gene | Primary Findings | COV | LCV |
| --- | --- | --- | --- |
| *AR* | Hub Gene, Drug Target, COVID-19 Severity | [15] | - |
| *ATOSA* | Downregulated in COVID-19 | [16] | - |
| *BTN3A1* | Predictive Marker | [17] | - |
| *CDKN1A* | Key Regulator, Drug Target | [18] | [19] |
| *CREBBP* | Hub/Drug Target | [20] | [21] |
| *EIF5A* | Drug Target | [22] | - |
| *EP300* | Hub/Drug/Vaccine Target, COVID-19 Sev., Epigenetic Reg. | [23] | [24] |
| *ESR1* | Hub/Drug Target, Herpes Zoster Association | [25] | - |
| *FYN* | Hub/Drug Target | [26] | - |
| *GRB2* | Drug Target | [27] | - |
| *HDAC1* | Drug Target, Epigenetic Regulation | [28] | [29] |
| *MAPK1* | Hub/Drug Target | [30] | - |
| *NDUFA6* | Drug Target | [31] | - |
| *RB1* | Hub Gene, SARS-CoV-2 Oncogenesis, Genetic Polymo. | [32] | [33] |
| *SMAD2* | Hub/Drug Target | [34] | [34] |
| *SMAD3* | Drug Target, Virus-host Interaction | [35] | - |
| *SRC* | Drug Target, Virus-host Interaction | [36] | [37] |
| *TP53* | Hub/Drug/Vaccine Target, Critical Gene | [38] | [39] |
| *YWHAG* | Hub/Vaccine Target, COVID-19 Neurotropism | [40] | - |

The remaining 13 genes in our causal gene set are novel discoveries in the context of COVID-19 and Long COVID research: adenosine deaminase tRNA-specific 1 (*ADAT1* ), B-cell lymphoma 2 interacting protein 1 (*BNIP1* ), bole-like 2 (*BOLA2* ), chromosome 19 open reading frame 18 (*C19orf18* ), inositol 1,4,5-trisphosphate receptor interacting domain containing 1 (*ITPRID1* ), *CDC26*, cytidine deaminase (*CDA*), ceramide synthase 4 (*CERS4* ), casein kinase 2 alpha 1 (*CSNK2A1* ), GDP-mannose pyrophosphorylase B synthase (*GMPPB* ), MORN repeat containing 3 (*MORN3* ), *MORN4* and the von Willebrand factor D and EGF domains gene (*VWDE* ). These genes have not been previously linked to COVID-19 or Long COVID, demonstrating the potential of our framework to reveal novel targets for intervention and further study.

Enrichment analysis of the 32 causal genes identified 458 significant pathways in GO (Gene Ontology) [41], 99 in KEGG (Kyoto Encyclopedia of Genes and Genomes) [42], and 246 in Reactome [43]. The top 20 pathways from each database, ranked by adjusted p-value, are shown in Fig. 3, with the complete list available in [Supplementary Data 3](https://github.com/SindyPin/Causal-Multiomics-Method/tree/main/SM/SM3).

**Fig. 3:**
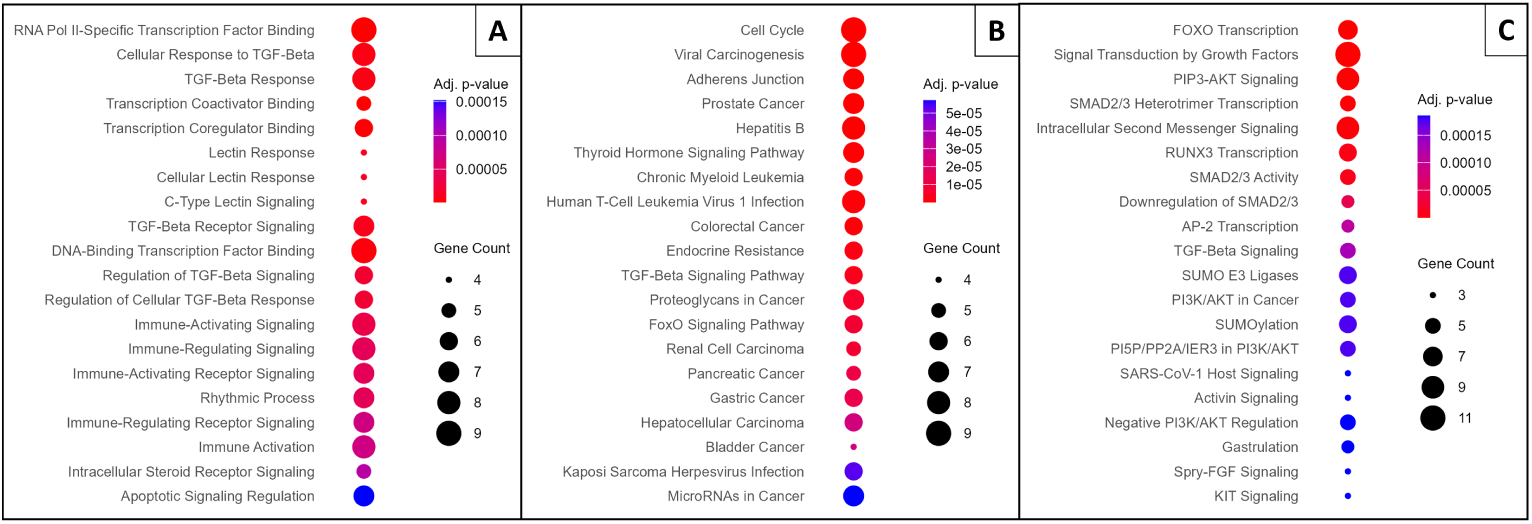
Enrichment analysis (EA) results for the identified Long COVID Causal Genes. (A) Gene Ontology (GO) EA, showing the top 20 enriched terms across Biological Process (BP) and Molecular Function (MF) categories. (B) KEGG pathway EA, displaying the top 20 enriched pathways. (C) Reactome pathway EA, illustrating the top 20 enriched pathways. For all plots, genes are ranked by the lowest adjusted p-value. The y-axis represents the enriched terms or pathways, the size of each dot reflects the number of associated genes, and the color gradient indicates the adjusted p-value, with red denoting greater significance.

Key findings include the transforming growth factor (TGF)-Beta signaling pathway, highlighted in GO and KEGG analyses, which plays a crucial role in immune regulation and tissue repair. Its disruption may contribute to persistent inflammation and fibrosis, leading to lung and organ damage as observed in Long COVID patients [18]. Similarly, KEGG pathways like cell cycle and viral carcinogenesis suggest long-term cellular effects of SARS-CoV-2 infection, such as abnormal proliferation and senescence, potentially explaining prolonged recovery and tissue dysfunction [33].

GO analysis highlights the importance of immune signaling pathways in ongoing inflammation and autoimmune-like symptoms [44]. The Reactome analysis emphasizes Forkhead box O (FOXO) transcription and Phosphoinositide 3-kinase (PI3K)/Protein Kinase B (AKT) signaling, which are involved in metabolism, stress responses, cell survival, and growth factor signaling pathways that may impair tissue repair and regeneration [18, 21].

These findings reveal potential mechanisms underlying Long COVID and suggest therapeutic targets, such as TGF-Beta signaling and FOXO transcription, to mitigate long-term effects.

#### 2.3.1 Shared Genetic Basis of Long COVID and Related Conditions

We examined the involvement of the 32 identified Long COVID causal genes in other pathophysiological conditions (Table 2, complete dataset in [Supplementary Data 4](https://github.com/SindyPin/Causal-Multiomics-Method/tree/main/SM/SM4)). This analysis revealed several distinct patterns of disease overlap, curated from multiple disease databases, including The Human Disease Database (MalaCards), Disease-Gene Associations (DISEASES), The Gene-Disease Network (DisGeNET), Medical Genetics Database (MedGen), and the Gene Curation Coalition (GenCC) (see the Methods section for more information about these databases). Many of these genes are implicated in a spectrum of syndromic, metabolic, autoimmune, connective tissue, and neurodevelopmental disorders that share clinical or biological features with Long COVID manifestations [7, 45, 46].

**Table 2:**
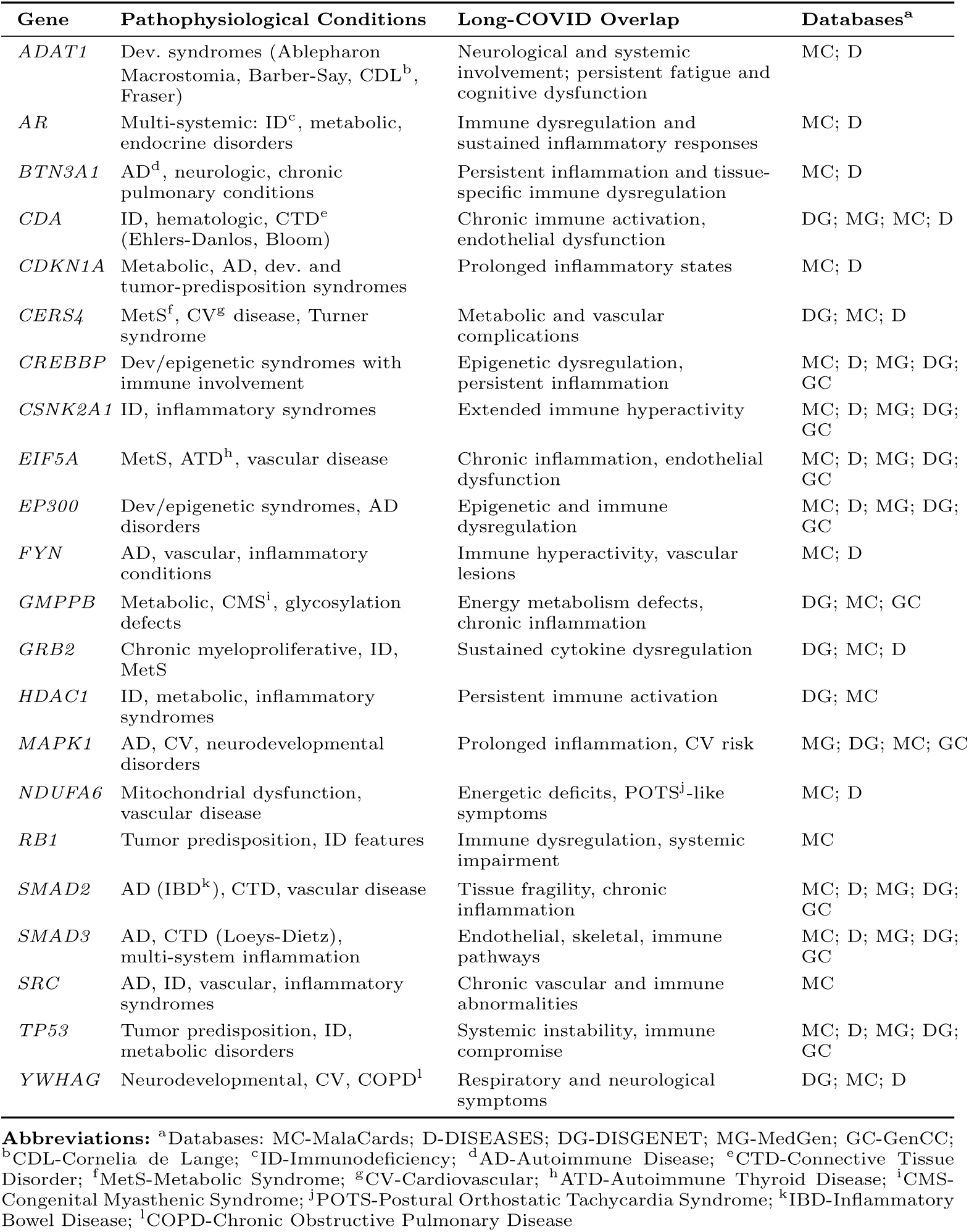
Causal genes in Long COVID and their overlap with other pathophysiological conditions. Analysis of the identified causal genes for Long COVID shows their involvement in other pathophysiological conditions and potential mechanistic overlap with Long COVID manifestations.

A subset of these genes (*CDKN1A*, *CREBBP*, *CSNK2A1*, and *TP53* ) are involved in tumor-predisposition syndromes and complex developmental disorders with autoimmune and inflammatory components. These conditions’ aberrant cytokine signaling and dysregulated immune checkpoints suggest potential mechanisms for prolonged inflammatory responses observed in Long COVID [7]. Similarly, genes such as *C19orf18*, *CDC26*, *MORN3*, *NDUFA6*, *VWDE*, and *YWHAG* are linked to systemic conditions affecting multiple organ systems. Their association with mitochondrial dysfunction and vascular pathologies parallels the fatigue, dysautonomia, and endothelial dysfunction commonly reported in Long COVID [45].

*ATOSA* and *GMPPB* are linked to chronic inflammation, mirroring mechanisms of immune activation and tissue damage implicated in Long COVID. Additionally, *CERS4*, *ESR1*, *FYN*, and *MAPK1* highlight the interplay between immune dysfunction and metabolic disruption, shedding light on the metabolic dysregulation seen in some patients [46].

Our database integration analysis reveals meaningful biological connections between Long COVID and other disorders, particularly immune-mediated conditions and metabolic diseases. The identified genetic overlaps suggest that variants in these genes might influence individual susceptibility to persistent post-viral symptoms, similar to their role in other chronic conditions. These shared molecular features help explain the diverse manifestations observed across Long COVID patients [47].

#### 2.3.2 Risk/Preventive Genes Driving Long COVID Susceptibility

Among the 32 causal genes obtained from our framework, we identified 16 significant protein-coding genes directly associated with the risk and prevention of Long COVID. These genes are implicated in critical biological processes such as cell cycle regulation (*CDC26* ), apoptosis (*BNIP1* ), and immune response (*BTN3A1* ) (Table 3).

**Table 3:**
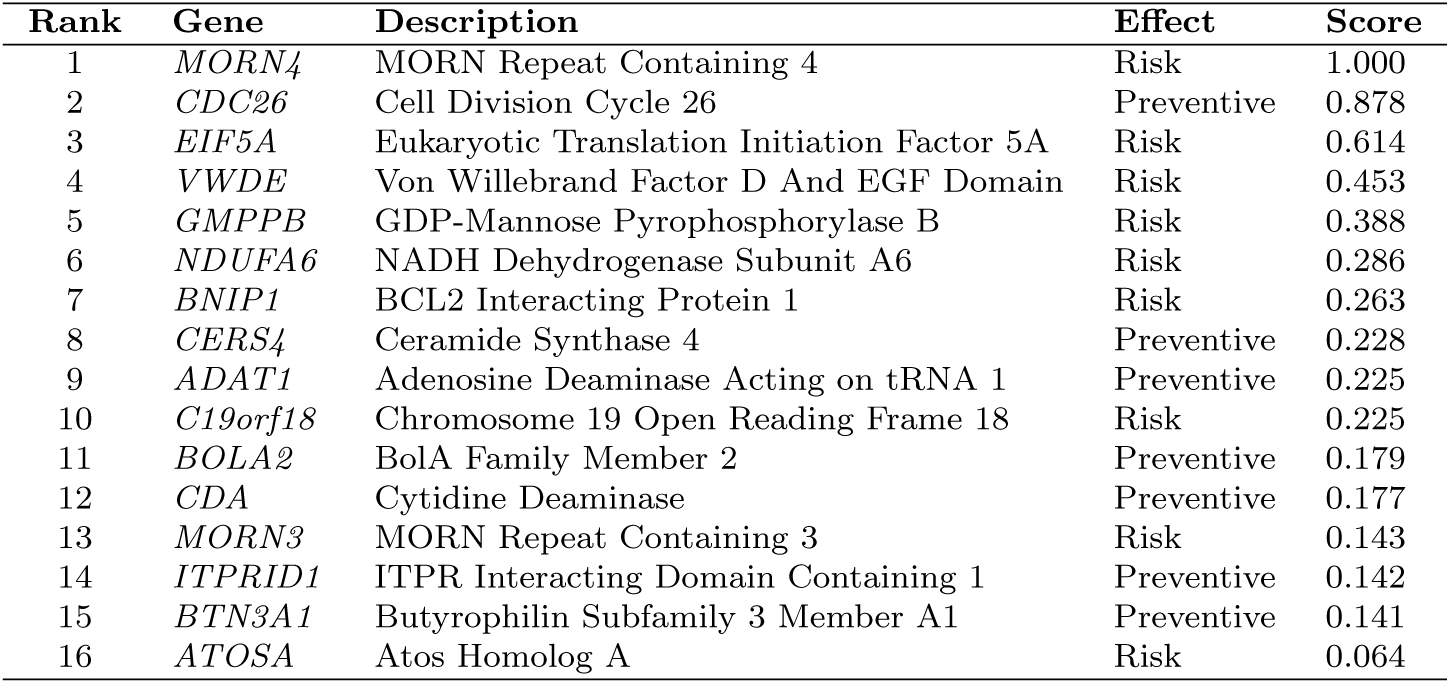
Risk and preventive causal genes for Long COVID ordered by the. *S***_Causal_ score.** Genes are classified as Risk or Preventive factors for Long COVID based on their effect size sign (positive or negative, respectively) when *α* = 1.

The forest plot (Fig. 4) reveals a wide range of effect sizes for 16 protein-coding genes, from -30.04 for *CDC26* to 34.22 for *MORN4*, indicating varying degrees of influence on Long COVID susceptibility. Applying our framework, we identified statistically significant causal relationships for these genes (p-value and False Discovery Rate (FDR) *<* 0.05), with confidence intervals that do not cross zero, providing strong evidence for their potential roles. Notably, genes such as *MORN4*, *CDC26*, *EIF5A*, and *VWDE* exhibit the strongest causal associations, with the largest absolute effect sizes.

**Fig. 4:**
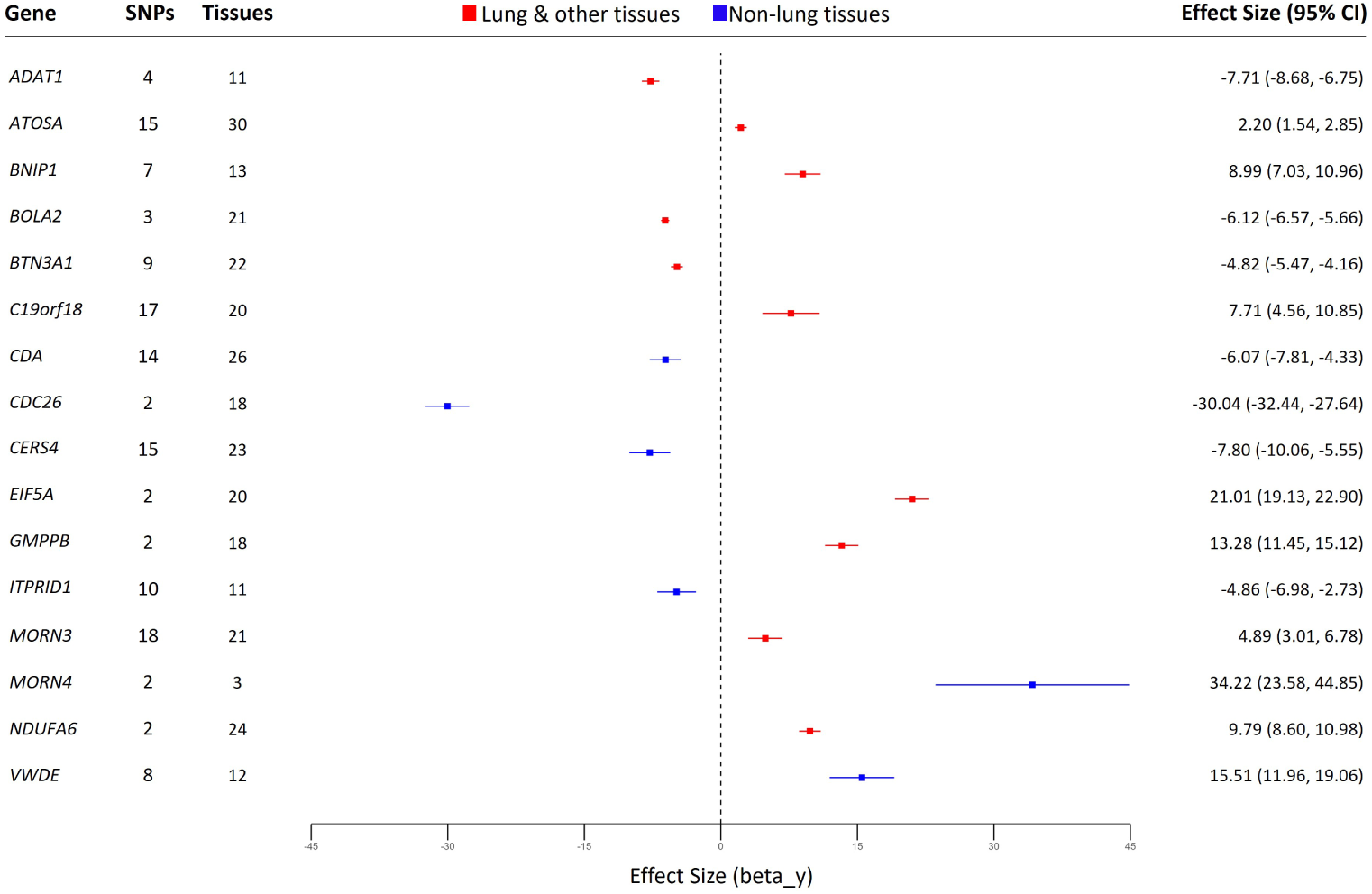
Effect size of the risk and preventive causal genes for Long COVID. Forest plot shows the significant genes identified at *α* = 1.0, with all causal relationships meeting statistical significance (p-value and FDR *<* 0.05). Higher expression is associated with increased (positive effect size) or decreased (negative effect size) risk. SNPs: number of associated SNPs; Tissues: number of tissues where the SNPs influence the gene expression. Points show fixed effect size (standardized beta coefficient) with 95% CI error bars. Red: lung and other tissues; Blue: non-lung tissues. GWAS: Genome-Wide Association Study. SNP: Single Nucleotide Polymorphism. FDR: False Discovery Rate. CI: Confidence Interval.

In our framework, we used varying numbers of SNPs as IVs for each gene’s expression—ranging from 2 SNPs for genes like in *MORN4*, *CDC26*, *EIF5A*, *GMPPB*, and *NDUFA6*, to 18 SNPs for *MORN3*. These IVs strengthen the validity of our causal estimates between gene expression and Long COVID risk. The number of the tissues in which the gene expression was evaluated also varied by gene, enhancing the robustness of our findings across different biological contexts. For instance, *MORN4* showed expression changes in two tissues/cells (left ventricle and thyroid) and in cultured fibroblasts. In contrast, *CDA* exhibited a broad impact, affecting gene expression across 26 distinct tissues. These span multiple systems: adipose, brain, cardiovascular, endocrine, connective, immune, digestive, reproductive, excretory tissues, and blood. This extensive tissue distribution highlights the far-reaching effects of SNPs on gene expression throughout the body.

Moreover, the expression patterns of all the 16 risk/preventive protein-coding genes identified through our framework suggest a systemic involvement in Long COVID. Ten genes showed expression in both lung and other tissues, while six genes were expressed exclusively in non-lung tissues. This distribution of expression patterns in other non-lung tissues supports the presence of non-respiratory symptoms observed in Long COVID patients, which suggests the involvement of molecular mechanisms beyond the pulmonary system [48].

The directional effects vary among the genes, with some showing positive effect sizes (e.g., *ATOSA*, *BNIP1*, *C19orf18*, *EIF5A*, *GMPPB*, *MORN3*, *MORN4*, *NDUFA6*, and *VWDE* ) and others negative effect sizes (e.g., *ADAT1*, *BOLA2*, *BTN3A1*, *CDA*, *CDC26*, *CERS4*, and *ITPRID1* ). Genes with positive effect sizes suggest increased expression in relevant tissues is associated with higher Long COVID susceptibility. In contrast, those with negative effect sizes indicate that increased expression may reduce the risk or be protective against Long COVID.

Among these 16 protein-coding genes, the roles of *BTN3A1*, *EIF5A*, and *NDUFA6* were previously identified in the pathogenesis of COVID-19, suggesting a potential link between their expression and the development of Long COVID. [17, 22, 31] (Table 4).

**Table 4:** Summary of three causal genes with established links to COVID-19 and hypothesized effects in Long COVID. COVID-19: Coronavirus Disease 2019. PRF: Programmed Ribosomal Frameshifting. Additional related literature and references can be found in the [Supplementary Data 2](https://github.com/SindyPin/Causal-Multiomics-Method/tree/main/SM/SM2).

| Gene | General Function | Role in COVID-19 | Long COVID Impact |
| --- | --- | --- | --- |
| *BTN3A1* | T-cell activation and regulation [51] | Part of 5-gene signature; higher expression correlates with more ventilator-free days [17] | Higher expression may reduce risk via improved immune regulation |
| *EIF5A* | Translation regulation, protein synthesis, virus response, cell differentiation/death [52] | Promotes PRF, translation termination and ribosome recycling in SARS-CoV-2 [22] | May contribute to persistent symptoms due to enhanced viral response |
| *NDUFA6* | NADH dehydrogenase activity, electron transport, energy production [52] | Top ten hub gene, significant mRNA differences [31] | Disruptions may increase risk by impacting cardiovascular health |

*BTN3A1*, an immune system protein involved in T-cell activation and regulation, is part of a 5-gene signature predicting ventilator-free days in COVID-19 patients [17]. Our analysis revealed a negative effect size value for *BTN3A1*, suggesting that higher expression is causally associated with better clinical outcomes and potentially reduced Long COVID risk. This protective effect may be attributed to its role in promoting a more controlled immune response, thereby reducing long-term complications [49].

Conversely, *EIF5A*, a translation factor that promotes programmed ribosomal frameshifting (PRF), translation termination, and ribosome recycling in SARS-CoV-2 infection, showed a positive effect size value. This function indicates that *EIF5A* could contribute to persistent symptoms in Long COVID through ongoing disruption in translation regulation and protein synthesis, resulting in continued immune activation and cellular stress [22].

*NDUFA6*, a key component of the mitochondrial respiratory chain, has been identified among the top genes associated with SARS-CoV-2 infection [31], showing significant mRNA expression differences in affected patients. Our findings indicate that disruptions in *NDUFA6* may adversely affect cardiovascular health and elevate Long COVID risk. These effects are likely attributable to the gene’s critical role in cellular energy production and mitochondrial function. Impaired activity of *NDUFA6* can lead to reduced ATP synthesis, increased oxidative stress, and the development of cardiovascular symptoms frequently observed in Long COVID patients [50].

These findings suggest that *BTN3A1*, *EIF5A*, and *NDUFA6* play significant roles in COVID-19 and may have implications in Long COVID, with *BTN3A1* potentially mitigating Long COVID risk through improved immune regulation, while *EIF5A* and *NDUFA6* contributing to persistent symptoms due to their roles in viral response and energy production, respectively.

In addition to the three previously mentioned genes, our framework identified 13 novel risk/preventive causal genes for Long COVID. Among these genes, *CDA*, *ADAT1*, *CERS4*, *CDC26*, and *BOLA2* were mainly enriched in our analyses with significant roles in crucial pathways, including nucleotide metabolism, RNA editing, lipid metabolism, cell cycle regulation, and iron-sulfur cluster assembly.

*CDA* and *ADAT1* are both involved in nucleotide metabolism and RNA editing processes. *CDA* is crucial for pyrimidine salvage and nucleotide pool balance, potentially impacting RNA integrity and immune system function [53]. Similarly, *ADAT1* is implicated in pre-mRNA editing, converting adenosine to inosine in eukaryotic tRNA, potentially influencing inflammatory responses [54]. Their roles as risk factors may be hypothesized based on their involvement in these critical cellular processes, which could contribute to the persistent symptoms observed in Long COVID patients [4].

*CERS4* and *BOLA2* are involved in cellular metabolism and homeostasis. *CERS4* facilitates sphingosine N-acyltransferase activity and is implicated in ceramide synthesis, influencing lipid metabolism and cellular signaling pathways [55]. *BOLA2* works in iron maturation and is part of the iron-sulfur cluster assembly complex, playing a role in cell redox homeostasis [56]. The risk association of these genes might be related to their impact on various cellular processes, including signaling pathways and cellular respiration. Its role may be associated with the diverse symptoms observed in Long COVID cases [57].

*CDC26* is part of the anaphase-promoting complex (APC) involved in cell cycle regulation [58]. Its role as a risk factor may be attributed to its function as a ubiquitin-protein ligase, managing the proteolysis of cell cycle proteins, which could impair cellular repair and regeneration processes, possibly explaining the prolonged cellular damage found in individuals with Long COVID [59].

Comprehensive results of pathway enrichment analyses using GO, KEGG, and Reactome databases, including significantly enriched biological processes, molecular functions, cellular components, and pathways, are detailed in [Supplementary Data 5](https://github.com/SindyPin/Causal-Multiomics-Method/tree/main/SM/SM5).

#### 2.3.3 Network Driver Genes Controlling Long COVID Network

There were 16 genes in the causal gene list found as network drivers of Long COVID. In the context of CT, network driver genes are critical nodes within a biological network whose manipulation can control the overall state and dynamics of the system. They serve as key regulators that can influence the activity of numerous downstream genes and pathways. Targeting these driver genes makes it possible to control the network toward a desired state, such as restoring normal function or mitigating disease effects. By identifying these genes, our framework highlights key intervention points within the network, offering potential targets to influence and modify its behavior and outcomes [13].

These identified core network driver genes have at least 150 connections with other nodes, emphasizing their significant influence. Disruption of these genes under normal conditions could contribute to the pathogenesis of Long COVID, making them potential therapeutic targets for restoring normal function in affected patients (Table 5).

**Table 5:**
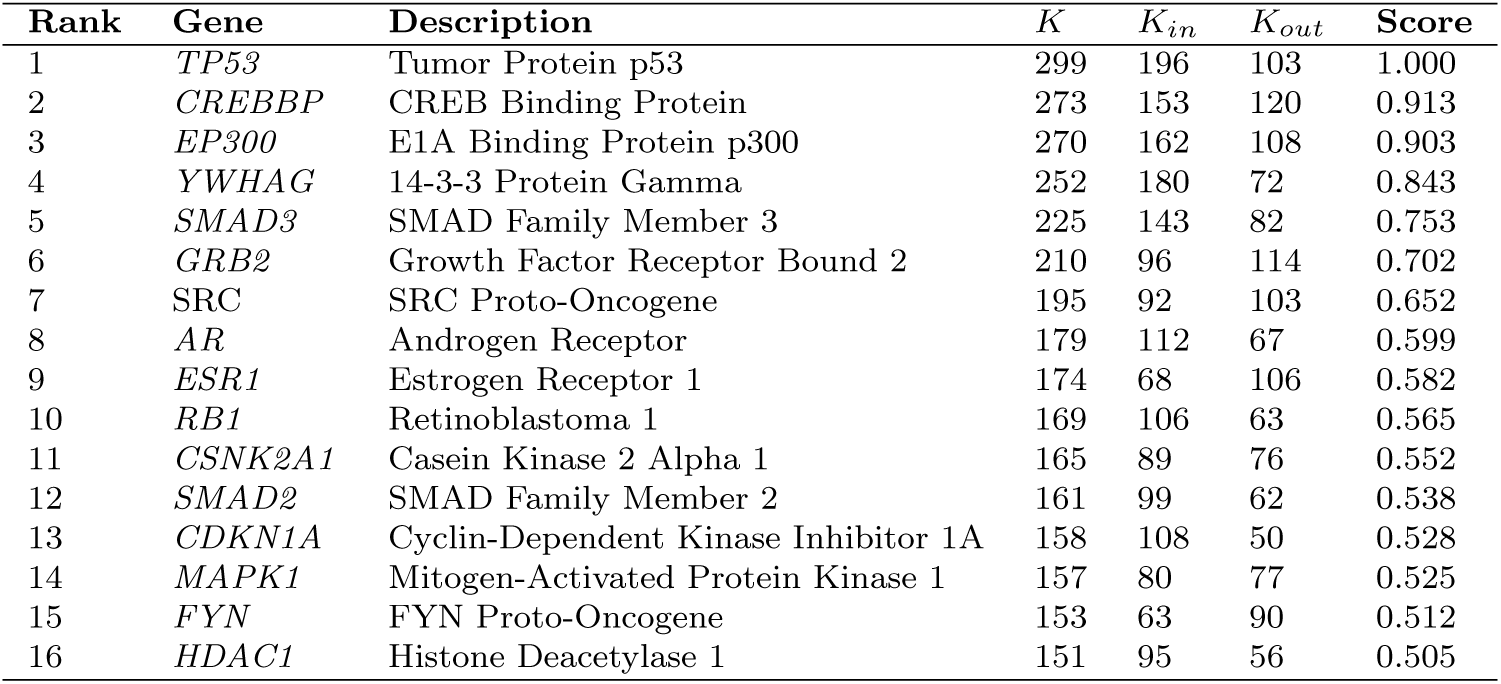
Network driver genes for Long COVID ordered by the. *S***_Causal_ score.** The K column represents the total degree (total number of interactions), *K_in_* describes the in-degree (incoming interactions), and *K_out_* denotes the out-degree (outgoing interactions).

| Rank | Gene | Description | $K$ | $K_{in}$ | $K_{out}$ | Score |
| --- | --- | --- | --- | --- | --- | --- |
| 1 | *TP53* | Tumor Protein p53 | 299 | 196 | 103 | 1.000 |
| 2 | *CREBBP* | CREB Binding Protein | 273 | 153 | 120 | 0.913 |
| 3 | *EP300* | E1A Binding Protein p300 | 270 | 162 | 108 | 0.903 |
| 4 | *YWHAG* | 14-3-3 Protein Gamma | 252 | 180 | 72 | 0.843 |
| 5 | *SMAD3* | SMAD Family Member 3 | 225 | 143 | 82 | 0.753 |
| 6 | *GRB2* | Growth Factor Receptor Bound 2 | 210 | 96 | 114 | 0.702 |
| 7 | *SRC* | SRC Proto-Oncogene | 195 | 92 | 103 | 0.652 |
| 8 | *AR* | Androgen Receptor | 179 | 112 | 67 | 0.599 |
| 9 | *ESR1* | Estrogen Receptor 1 | 174 | 68 | 106 | 0.582 |
| 10 | *RB1* | Retinoblastoma 1 | 169 | 106 | 63 | 0.565 |
| 11 | *CSNK2A1* | Casein Kinase 2 Alpha 1 | 165 | 89 | 76 | 0.552 |
| 12 | *SMAD2* | SMAD Family Member 2 | 161 | 99 | 62 | 0.538 |
| 13 | *CDKN1A* | Cyclin-Dependent Kinase Inhibitor 1A | 158 | 108 | 50 | 0.528 |
| 14 | *MAPK1* | Mitogen-Activated Protein Kinase 1 | 157 | 80 | 77 | 0.525 |
| 15 | *FYN* | FYN Proto-Oncogene | 153 | 63 | 90 | 0.512 |
| 16 | *HDAC1* | Histone Deacetylase 1 | 151 | 95 | 56 | 0.505 |

Of the 16 network driver genes identified, 14 were associated with enriched pathways crucial in COVID-19, Long COVID, or both. These pathways involved essential cellular functions such as cell proliferation, differentiation, cycle progression, DNA repair, inflammation, and immune responses. Disruptions in these processes can lead to the persistent symptoms of Long COVID, chronic inflammation, neurodegeneration, and immune dysfunction (Table 6).

**Table 6:** Long COVID roles of the identified network driver genes. Key protein functions and enriched pathways obtained from GO, KEGG, or Reactome are shown, along with their roles in COVID-19 and Long COVID pathogenesis, illustrating their contribution to disease mechanisms. All pathway enrichments meet statistical significance thresholds (p-value and FDR *<* 0.05). FDR: False Discovery Rate. Additional references can be found in the [Supplementary Data 2](https://github.com/SindyPin/Causal-Multiomics-Method/tree/main/SM/SM2).

| Gene | Function | Paths | Main Path | Roles | Ref. |
| --- | --- | --- | --- | --- | --- |
| *AR* | Steroid-hormone transcription factor, regulates cell proliferation | 88 | Regulation of miRNA transcription | Affects TMPRSS2 and ACE2 expression, linked to persistent symptoms in males | [60] |
| *CDKN1A* | Inhibits CDKs, regulates cell cycle | 152 | p53 signaling pathway | Involved in SARS-CoV-2 entry, tissue damage, fibrosis | [61] |
| *CREBBP* | Acetyltransferase, regulates gene expression | 118 | Histone acetyltransferase activity | Controls inflammation, may trigger neurodegeneration | [21] |
| *EP300* | Acetyltransferase, regulates cell growth | 184 | Histone acetyltransferase activity | Regulates ACE2, critical in inflammation, persistent immune responses | [24] |
| *ESR1* | Estrogen receptor, regulates transcription | 113 | Intracellular estrogen receptor signaling pathway | Protective against COVID-19, reduces inflammation, immune dysfunction in women | [62] |
| *FYN* | Non-receptor kinase, regulates immune response | 115 | Immune response-regulating signaling pathway | Regulates inflammation, may be linked to immune dysregulation | [44] |
| *HDAC1* | Histone deacetylase, modulates gene expression | 98 | Regulation of apoptotic signaling pathway | Modulates inflammation and apoptosis in COVID-19 | [63] |
| *MAPK1* | Kinase involved in signal transduction | 246 | Immune response-activating signaling pathway | Controls inflammation and cytokine responses in COVID-19 | [64] |
| *RB1* | Tumor suppressor, regulates cell cycle | 25 | Regulation of apoptotic signaling pathway | May interact with viral mechanisms, potential oncogenic effects | [33] |
| *SMAD2* | Mediates TGF-beta signals, regulates cell growth | 114 | Transforming growth factor beta receptor signaling pathway | Involved in fibrosis and other complications post-COVID | [61] |
| *SMAD3* | Mediates TGF-beta signals, regulates cell differentiation | 168 | miRNA transcription | Linked to pulmonary fibrosis, impacts post-COVID severity | [61] |
| *SRC* | Non-receptor kinase, regulates gene transcription | 257 | Immune response-regulating signaling pathway | Mediates viral entry, chronic inflammation, immune dysregulation | [37] |
| *TP53* | Tumor suppressor, regulates apoptosis and DNA repair | 263 | Intrinsic apoptotic signaling pathway in response to DNA damage | Influences cytokine release, immune response in COVID-19 | [65] |
| *YWHAG* | Adapter protein in signaling pathways | 26 | PI3K-Akt signaling pathway | Involved in cell survival, inflammation, and immune responses in COVID-19 | [66] |

Extensive findings from our functional enrichment studies on these network driver causal genes, obtained from GO, KEGG, and Reactome, are presented in detail in [Supplementary Data 6](https://github.com/SindyPin/Causal-Multiomics-Method/tree/main/SM/SM6).

In Fig. 5, we provide a detailed example of *CREBBP*, one of the genes identified by our framework and confirmed in the literature. This gene was chosen due to its higher number of connections compared to other genes, highlighting its essential role in the network. The plots of the other identified network driver protein-coding genes for Long COVID are provided in the [Supplementary Data 7](https://github.com/SindyPin/Causal-Multiomics-Method/tree/main/SM/SM7).

**Fig. 5:**
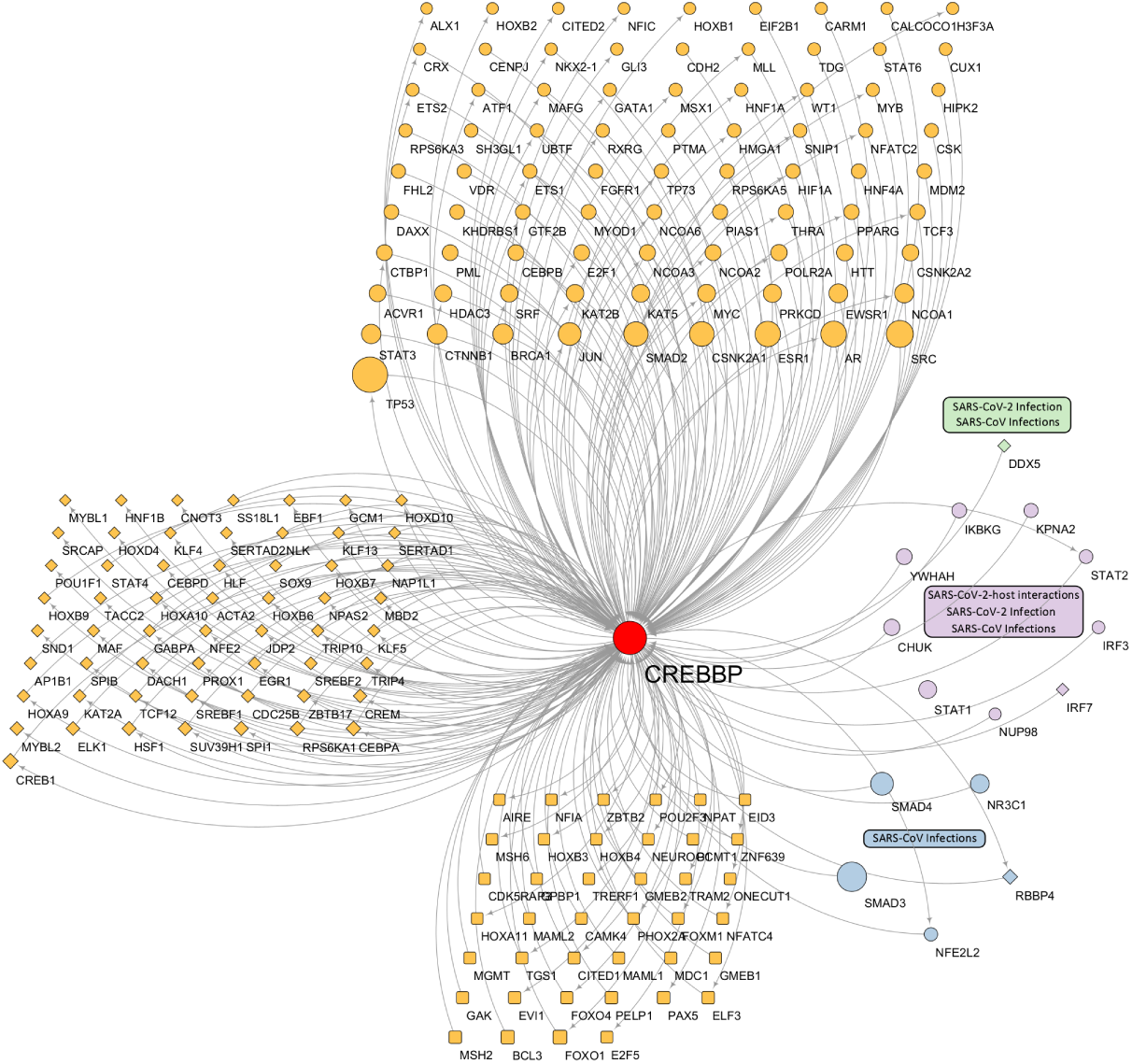
Network plot highlighting a network driver gene for Long COVID. Our analysis identified *CREBBP* as a key network driver gene, supported by existing literature, with 273 total interactions (153 incoming, 120 outgoing). Connected genes are represented by three shapes based on network control properties: ellipses for critical genes (whose absence increases required driver nodes), diamonds for ordinary genes (whose removal maintains current driver nodes), and round rectangles for redundant genes (whose removal preserves network control). The three most enriched pathways are shown in green, purple, and blue, with node sizes proportional to their K-degree (network connectivity).

### 2.4 Gene Expression Clustering Reveals Long COVID Subtypes

We clustered Long COVID patients into subgroups using gene expression data from the 32 identified causal genes. Moreover, we hypothesized that distinct gene expression patterns of risk/preventative genes and network driver genes underlay different clinical characteristics in patients. Using Consensus Clustering (ConC) [67], we identified patient subgroups that demonstrated coherent clustering and balanced distributions (i.e., not skewed toward a single subset).

The analysis identified three distinct Long COVID subtypes, aligning with the three-cluster findings reported in previous research [4, 45], each with high Average Silhouette Width (ASW) values indicating robust clustering: Cluster 1 included 65 individuals (ASW: 0.93), Cluster 2 contained 53 individuals (ASW: 0.85), and Cluster 3 consisted of 36 individuals (ASW: 0.75).

As shown in Fig. 6, these causal genes exhibited distinct expression patterns across subtypes, highlighting their potential role in differentiating symptom profiles. To explore this further, we mapped symptom prevalence within clusters to evaluate whether gene expression patterns reliably aligned with the identified symptoms. Significant differences in symptom distributions (p-value *<* 0.05) were observed, with symptoms grouped into broader categories, including respiratory, gastrointestinal, neurological, metabolic, psychological, dental, and sleep-related issues, enabling a comprehensive comparison across clusters. Table 7 complements this analysis by summarizing the key genes identified per cluster, their regulation patterns, biological functions, and associated enriched pathways contributing to the distinct Long COVID symptom manifestations [68]. Details of the RNA-seq and clinical datasets used in this analysis are provided in the Methods section.

**Fig. 6:**
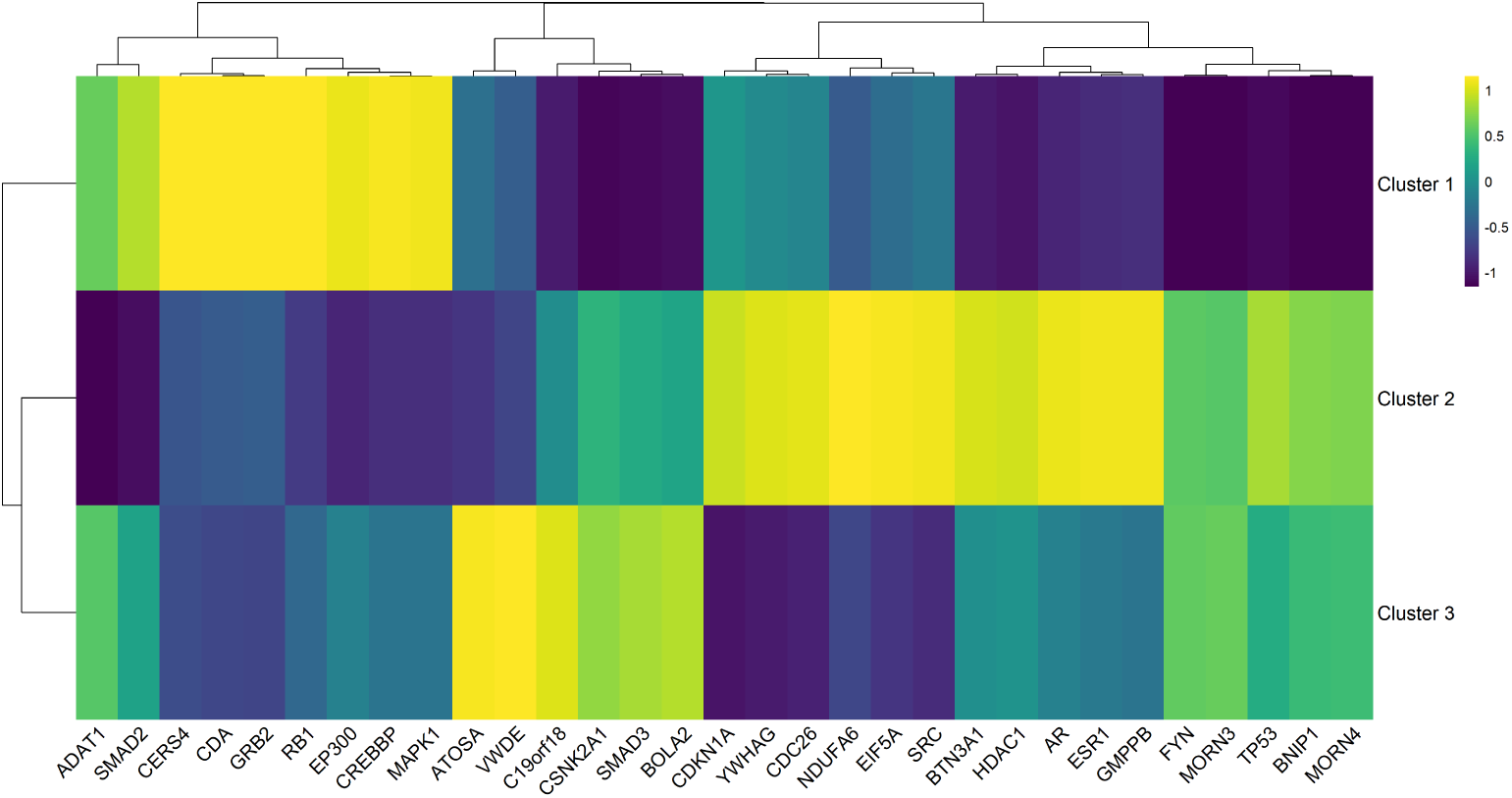
Gene expression heatmap with hierarchical clustering of Long COVID patients revealing three distinct clusters. Normalized expression values range from -1 (dark purple, low expression) to 1 (bright yellow, high expression) across identified genes and patient clusters. Dendrograms on both axes show the hierarchical relationships between genes (top) and clusters (left), with height indicating the degree of similarity between clusters. Gene names are displayed on the x-axis, and cluster identifiers are shown on the y-axis.

**Table 7:** Gene expression patterns, pathways, and symptoms across Long COVID clusters. Cluster-specific genes highlight functions and enriched pathways associated with symptom persistence. The table shows relationships between clusters, symptoms, and pathways with significant biological relevance (p-values and FDR *<* 0.05). Clus.: Cluster. Reg.: Regulation. FDR: False Discovery Rate

| Clus | Symptoms | Gene | Reg. | Function | Enriched Pathway |
| --- | --- | --- | --- | --- | --- |
| 1 | Respiratory issues,<br>Sleep disturbances | *CREBBP* | Up | Transcriptional coactivator, Hypoxia response | HIF-1 signaling: mediates cellular response to hypoxia |
|  |  | *GRB2* | Up | Growth factor signaling mediator | ErbB signaling: regulates cell survival and stress response |
|  |  | *MAPK1* | Up | Stress-responsive kinase | MAPK signaling: controls cellular response to stress and inflammation |
|  |  | *SMAD2* | Up | Signal transducer in TGF-beta pathway, Regulates inflammation | TGF-beta signaling: controls inflammatory response and tissue repair |
| 2 | Psychological symptoms, Dental issues | *CDC26* | Up | Cell cycle regulator | Controls cellular homeostasis |
|  |  | *CDKN1A* | Up | Cell cycle regulator, Stress response | p53 signaling: mediates cellular stress response |
|  |  | *ESR1* | Up | Nuclear receptor, Inflammation control | Nuclear receptor signaling: regulates inflammatory responses |
|  |  | *YWHAG* | Up | Signal transduction regulator | PI3K-Akt signaling: controls cell survival and stress adaptation |
| 3 | Gastrointestinal symptoms, Metabolic disturbances | *HDAC1* | Down | Epigenetic regulator | Chromatin modification: regulates gene expression |
|  |  | *NDUFA6* | Up | Mitochondrial function | Oxidative phosphorylation: controls energy metabolism |
|  |  | *SRC* | Down | Tyrosine kinase, Immune regulation | Immune response signaling: controls inflammation |
|  |  | *TP53* | Down | Stress response regulator | Apoptotic signaling: regulates cell death and survival |

Cluster 1 showed a symptom profile dominated by respiratory issues and sleep disturbances. Increased mucus was reported by 29.23% of patients in this cluster, significantly higher than in cluster 2 (15.09%) and cluster 3 (16.67%) (*χ*^2^ *p* = 1.07*×*10*^−^*^41^, adjusted *p* = 1.18 *×* 10*^−^*^40^). Lung problems (23.08%) and smell and/or taste problems (20.00%) were similarly more prevalent in cluster 1 (*χ*^2^ *p* = 7.69 *×* 10*^−^*^7^, adjusted *p* = 1.21 *×* 10*^−^*^6^; *χ*^2^ *p* = 8.23 *×* 10*^−^*^15^, adjusted *p* = 2.59 *×* 10*^−^*^14^, respectively). Sleep problems were also more common in cluster 1 (49.23%) compared to cluster 2 (28.30%) and cluster 3 (33.33%) (*χ*^2^ *p* = 9.98 *×* 10*^−^*^33^, adjusted *p* = 5.49 *×* 10*^−^*^32^), aligning with previous reports indicating sleep disturbances as key features of specific Long COVID phenotypes [69]. This pattern is consistent with multiple cluster analyses identifying distinct respiratory and fatigue-related symptom groups [4, 45]. The corresponding gene expression profile showed elevated expression of *CREBBP*, *GRB2*, *MAPK1*, and *SMAD2*, which are involved in inflammatory responses, stress adaptation, and TGF-beta signaling pathways linked to respiratory function and sleep regulation. These molecular findings suggest that the selected genes in cluster 1 effectively capture the biological mechanisms underlying this group’s respiratory and sleep-related symptoms.

A higher prevalence of psychological symptoms and dental issues characterized cluster 2. Anxiety and depression were observed in 37.74% of patients, slightly higher than in cluster 1 (36.92%) and significantly higher than in cluster 3 (25.00%) (*χ*^2^ *p* = 0.0082, adjusted *p* = 0.0106). Cavities and teeth problems affected 18.87% of cluster 2 patients, compared to 13.85% in cluster 1 and 5.56% in cluster 3 (*χ*^2^ *p* = 2.32 *×* 10*^−^*^9^, adjusted *p* = 4.65 *×* 10*^−^*^9^). Gene expression analysis in cluster 2 revealed upregulation of *CDC26*, *CDKN1A*, *ESR1*, and *YWHAG*, genes associated with cell cycle regulation, stress response, and inflammation control, respectively. Notably, *ESR1* has been implicated in psychiatric disorders, and *YWHAG* is known to modulate multiple signaling pathways relevant to mood regulation [70]. The prominence of neuropsychological symptoms in this cluster aligns with other Long COVID clustering studies that have identified distinct neurocognitive and mood-related phenotypes [4, 45]. Furthermore, recent studies suggest an interplay between COVID-19 and oral health deterioration, providing a rationale for the increased dental issues in cluster 2 [71]. These findings reflect this cluster’s biological mechanisms tied to psychological and dental symptoms.

Cluster 3 was defined by gastrointestinal (GI) symptoms and metabolic disturbances. A significant 38.89% of patients in this cluster experienced nausea, diarrhea, and/or vomiting, higher than in cluster 1 (13.85%) and cluster 2 (7.55%) (*χ*^2^ *p* = 1.37*×*10*^−^*^116^, adjusted *p* = 3.02*×*10*^−^*^115^). Eating more or less was reported by 47.22% of patients in cluster 3, comparable to cluster 1 (47.69%) but higher than cluster 2 (37.74%) (*χ*^2^ *p* = 1.56 *×* 10*^−^*^13^, adjusted *p* = 4.29 *×* 10*^−^*^13^). Headaches were also more common in cluster 3 (33.33%) compared to cluster 1 (30.77%) and cluster 2 (22.64%) (*χ*^2^ *p* = 2.19 *×* 10*^−^*^20^, adjusted *p* = 8.02 *×* 10*^−^*^20^). The gene expression profile showed downregulation of *HDAC1*, *SRC*, and *TP53*, along with upregulation of *NDUFA6*, genes associated with metabolic regulation, immune response, and cellular stress pathways. These alterations correlate with evidence of persistent metabolic and immune dysregulation in Long COVID [46]. The prominent GI issues are consistent with the recognition of GI-focused Long COVID clusters [4, 45], showing the heterogeneous nature of post-COVID symptom profiles. These molecular profiles correlate with the GI and metabolic symptoms identified in cluster 3, highlighting the ability of these genes to capture the biological processes driving these manifestations.

Integrating symptom profiles with gene expression clustering demonstrates how our identified genes stratify Long COVID patients into biologically distinct groups, each cluster exhibiting unique symptom signatures. Cluster 1 shows predominantly respiratory and sleep disturbances, suggesting potential benefits from therapies targeting respiratory and sleep pathways. Cluster 2 features psychological and dental issues, indicating the need for interventions addressing stress-related pathways and oral health. Cluster 3 presents GI and metabolic symptoms, suggesting treatments focused on metabolic and digestive support. The alignment between gene functions and symptom distributions validates the biological relevance of these causal genes and their role in driving diverse clinical manifestations. Further details, including complete statistical analyses and p-values, are available in [Supplementary Data 8](https://github.com/SindyPin/Causal-Multiomics-Method/tree/main/SM/SM8).

## 3 Discussion and Conclusion

Long COVID, or PASC, is characterized by the persistence or development of symptoms as a result of SARS-CoV-2 infection, affecting multiple systems, including respiratory, neurological, and cardiovascular systems [1–5]. Despite the increasing recognition of Long COVID, the causal genetic factors contributing to this condition remain primarily unidentified, posing a significant barrier to developing effective diagnostic and therapeutic strategies. Our work addressed this gap by introducing a novel multi-omics framework that integrated genetics, transcriptomics, and proteomics data to identify risk and preventive causal genes and network driver genes for Long COVID, ultimately guiding the development of targeted interventions. This approach enhances our understanding of Long COVID pathophysiology and provides a robust platform for further research into this disease.

Previous studies have highlighted the complexity and heterogeneity of Long COVID, identifying various risk factors and biomarkers related to inflammation and immune function [7]. However, these observational studies have not established a causal relationship between genes and Long COVID. In contrast, our computational analysis identified several significant causal genes for Long COVID, supported by existing literature and novel candidates to direct future work. These genes are implicated in key biological processes potentially explaining the persistent symptoms observed in Long COVID cases. EA further highlighted pathways related to SARS-CoV-2 response, viral carcinogenesis, cell cycle regulation, and immune system processes, providing deeper insights into the pathophysiological mechanisms of Long COVID.

Our analysis also demonstrated the utility of gene expression-based clustering in stratifying Long COVID patients into biologically meaningful subgroups, each with distinct symptom profiles and underlying molecular mechanisms. Cluster 1 exhibited respiratory and sleep disturbances linked to genes involved in inflammatory responses and TGF-beta signaling. Cluster 2 was characterized by psychological symptoms and dental issues associated with genes regulating stress response and cellular homeostasis. Cluster 3 showed GI and metabolic symptoms driven by genes related to metabolic regulation and digest pathways. These findings demonstrate the heterogeneity of Long COVID and highlight the potential for specific therapeutic interventions based on each cluster’s molecular and clinical characteristics. Future research should focus on validating these clusters and exploring personalized treatment strategies.

Comparison with existing studies reveals that our integrated approach offers a comprehensive understanding of Long COVID’s genetic basis, providing robust causal evidence in the Long COVID network. Given that the genetics of Long COVID remain largely unknown, our framework uses the parameter *α* as an exploratory tool to discover potential causal genes from different analytical perspectives. We designed *α* as an adjustable parameter that allows researchers to investigate dynamically the interplay between MR and CT. To facilitate broader research and enable the discovery of new potential causal genes across different populations, we have open-sourced our framework and developed a user-friendly Shiny app (https://sindypin.shinyapps.io/github/). This allows researchers to analyze their own datasets, explore different configurations, and reproduce our results.

In particular, as *α* decreases, our model revealed potential network driver genes, highlighting factors such as *TP53*, *CREBBP*, *EP300*, *YWHAG*, *SMAD3*, and *GRB2* that may play crucial roles in network control. Our framework identified these candidate genes from the studied populations, providing initial insights into Long COVID genetics. This approach can be applied to emerging genetic studies, enabling both the validation of current findings and the discovery of new causal genes across diverse populations. This iterative discovery process will help build a more comprehensive understanding of the Long COVID’s genetic architecture.

Our framework enables several future directions. As clinical and experimental studies establish the causal genes of Long COVID, researchers will be able to refine the *α* parameter for predictive applications. Additionally, integrating our approach with new data types, such as single-cell RNA-seq (scRNA-seq), could reveal more profound insights into cell-specific gene expression changes and heterogeneity. This could help identify how different cell types contribute to the development and persistence of Long COVID symptoms, potentially leading to more targeted therapeutic strategies.

Our analysis of the causal genes’ involvement in other pathophysiological conditions revealed important connections that support and extend previous findings [7, 47]. The overlap with conditions characterized by immune dysregulation, chronic inflammation, and multi-system involvement aligns with observed Long COVID manifestations [45, 46]. Identifying genes involved in developmental disorders and inflammatory conditions is crucial, suggesting that genetic predisposition to certain disorders might influence Long COVID susceptibility [7]. This connection between other conditions and Long COVID provides additional context for understanding disease heterogeneity and could inform risk assessment strategies [47]. The database integration approach also highlights how existing knowledge of genetic disorders can inform our understanding of novel post-viral conditions, demonstrating the value of using established disease databases in new disease research.

Beyond these immediate directions, the core set of 32 causal genes identified through our approach holds broad potential for clinical translation. Diagnostic kits using molecular assays (e.g., targeted polymerase chain reaction panels or multiplex protein-based tests) could be developed to detect abnormal expression patterns of these key genes, offering a more objective and standardized approach to diagnosing Long COVID. Integrating these biomarkers into machine learning models may further refine patient stratification, allowing reliable prediction of who is at higher risk and guiding early interventions. Such predictive models, trained on independent cohorts and incorporating multi-omic signatures, could ultimately be adapted into clinical decision support tools, helping healthcare professionals identify at-risk individuals before severe or persistent symptoms arise.

In addition, these core causal genes may reveal novel therapeutic paths. Researchers can conduct targeted drug screening by identifying driver genes in the underlying molecular network or consider repurposing existing therapeutics. Interventions that modulate the activity or expression of these genes could potentially reduce Long COVID symptoms, complementing supportive care with more precise treatments. To realize these possibilities, further validation in *in vitro* and *in vivo* models and longitudinal studies are essential to assess whether modulating these genes influences patient outcomes over time. As data on cellular heterogeneity expands through scRNA-seq and other advanced profiling techniques, these causal genes can serve as molecular markers, guiding the search for cell-type-specific interventions and personalized medicine strategies.

In conclusion, our study establishes a robust framework for identifying causal genes for Long COVID, providing critical insights into its pathophysiology and supporting the development of targeted therapeutic strategies. By integrating the discovered biomarkers into diagnostic assays, predictive modeling, and experimental therapeutics, we move closer to actionable clinical applications. Together, these findings have the potential to significantly improve the management and outcomes of individuals affected by Long COVID, offering a path toward more precise, evidence-driven care.

## 4 Methods

### 4.1 Input Data Collection and Preparation

The success of our integrative multi-omics framework relies on the careful selection and preparation of diverse datasets that capture Long COVID’s genetic, transcriptomic, and proteomic dimensions. We collected and curated high-quality data from publicly available resources, ensuring robust coverage of key biological processes. These datasets include cis-eQTL information from the Genotype-Tissue Expression (GTEx) project [72], GWAS findings for Long COVID susceptibility [73], Whole Genome Sequencing (WGS) for Linkage Disequilibrium (LD) analysis [72], and gene-level data from Ensembl [74]. Additionally, RNA-seq [75], and PPI network datasets were incorporated to provide a comprehensive view of gene expression and functional interactions. The following subsections detail the sources, characteristics, and preparation steps for each dataset used in our analysis.

#### 4.1.1 Expression Quantitative Trait Loci (eQTL)

We utilized 49 significant cis-eQTL datasets, each within a 1Mb region and meeting an FDR threshold of *<* 0.05, obtained from the GTEx project (Version 8, Ensembl 99, GRCh38) [72]. These datasets contain 39,832 unique genes derived from nearly 1,000 healthy European individuals, accessed on 09 August 2023. They were crucial for investigating the relationship between genetic variation and gene expression across different human tissues ([Supplementary Data 9](https://github.com/SindyPin/Causal-Multiomics-Method/tree/main/SM/SM9)). For more details and a description of the datasets available in the GTEx consortium, refer to the original publication [76].

#### 4.1.2 Genome-Wide Association Studies (GWAS)

We sourced a Long COVID GWAS dataset (Release 7; Ensembl 109; HGB GRCh38) from the study by Lammi et al., 2023 [73]. This dataset consists of 3,018 cases and 1,093,995 controls from six different ancestries, all of whom were evaluated for 19 symptoms three months post-COVID-19 infection according to the WHO and CDC definitions of Long COVID [1, 2]. For comprehensive details, including the complete list of ancestries, symptoms, and unique SNPs, please refer to the detailed tables and figures in [Supplementary Data 9](https://github.com/SindyPin/Causal-Multiomics-Method/tree/main/SM/SM9).

#### 4.1.3 Whole Genome Sequencing (WGS)

To ensure the robustness and validity of our method, we calculated the LD matrix retrieving the WGS BAM files from GTEx (Ensembl 88, GRCh38) that contains 820,792 unique SNPs from 836 European male and female individuals ([Supplementary Data 9](https://github.com/SindyPin/Causal-Multiomics-Method/tree/main/SM/SM9)). Access to this specific dataset was granted through special permission [72].

#### 4.1.4 Human Genes Dataset

To assess each gene’s causal relationship with the outcome, we utilized the public Human Genes dataset from the Ensembl Genes database (version 110, GRCh38), which contains 70,116 genes [74].

#### 4.1.5 RNA Sequencing (RNA-seq)

Moreover, we analyzed RNA-seq gene expression data from the Mount Sinai COVID-19 Biobank Study [68]. The dataset comprises patients with Long COVID symptoms (persisting for more than one-month post-acute infection, following established institutional criteria [3–5]), COVID-19 patients, and healthy controls. We sourced this dataset from the Gene Expression Omnibus - National Center for Biotechnology Information (GEO - NCBI) database under the identifier GSE215865, corresponding to the Ensembl GRCh37 release [75]. It contains 413 blood samples from 158 Long COVID individuals ([Supplementary Data 9](https://github.com/SindyPin/Causal-Multiomics-Method/tree/main/SM/SM9)).

#### 4.1.6 Protein-Protein Interaction (PPI)

Finally, we employed the human PPI dataset published by Vinayagam et al., 2011 [77] as a model for building the Long COVID network ([Supplementary Data 9](https://github.com/SindyPin/Causal-Multiomics-Method/tree/main/SM/SM9)).

### 4.2 Framework

To create a comprehensive list of causal genes for Long COVID and understand their roles in regulating the disease, we used a fusion approach that integrated MR and CT. Specifically, we calculated the scores of each gene using the formula in Equation 1. This approach produced a final ranking of genes based on their direct causal relationships and significance within the Long COVID network. The following sections detail the calculations of *S*_Risk_ and *S*_Network_

#### 4.2.1 Calculating *S*_Risk_

To calculate *S*_Risk_, we employed the *Mt-Robin* method [10] to identify genes acting as risk or protective factors for Long COVID. Using GWAS (**D**_GWAS_) and eQTL (**D**_eQTL_) data (Section 4.1), this approach accurately infers the dependence (fixed effects) between eQTL and GWAS effects, even with invalid IVs.

We first constructed and refined the LD matrix using SNPs from our dataset to ensure robust genetic instruments. We calculated pairwise *r*^2^ values and applied an LD 0.5 threshold to filter highly linked SNPs. Our multi-criteria SNP selection process eliminated those with multiple correlations above the LD threshold, prioritized SNPs present across multiple tissues with consistent effect directions, and selected significant SNPs with the smallest minimum p-values. Additionally, we required genes to be expressed in at least one tissue.

The statistical analysis involved reverse regression coefficients and weighted regression with random slopes and correlated errors. We integrated these results with GWAS standard errors and the refined LD matrix to inform our resampling strategy. We evaluated causal relationships through bootstrapping by generating null distributions while preserving the SNP LD structure. We resampled GWAS effect sizes for each gene, maintaining LD correlations, and calculated test statistics under the null hypothesis of no association. The p-value for each gene was determined by the proportion of null test statistics exceeding the observed value, excluding samples with non-convergence or singular fits in the mixed-effects model.

The final score calculation used the absolute effect size (*β_y_*) from the MR method. Genes with a p-value or FDR exceeding 0.05 received a score of 0, ensuring only significant causal effects. We normalized the MR score (*S*_Risk_) using min-max scaling for cross-gene comparability:

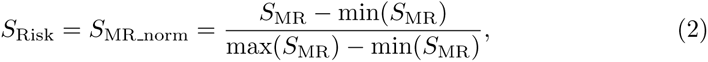

where *S*_MR_ represents each gene’s causal effect size, and min(*S*_MR_) and max(*S*_MR_) are the smallest and largest values across all genes, respectively.

Finally, we estimated FDR-corrected p-values to identify statistically significant causal contributors to Long COVID (p-value *<* 0.05).

#### 4.2.2 Calculating *S*_Network_

To calculate *S*_Network_, we analyzed gene impact using Long COVID RNA-seq expression data and the human PPI network (Section 4.1). Driver nodes are specific genes or proteins that, when manipulated, can control the state of the entire network. Network control refers to regulating or stabilizing the biological network by targeting these critical nodes. We first classified genes by removing each from the network and observing changes in the number of required driver nodes needed for control [13]. This process identified three categories: indispensable genes (increase in driver nodes needed), neutral genes (no significant change), and dispensable genes (minimal impact). We focused our analyses on indispensable genes due to their critical role in maintaining network control.

We further refined indispensable genes into Type-I and Type-II classifications based on their network behavior. Type-I genes were categorized by their effect on other driver nodes. Critical genes were those whose removal increases the number of required driver nodes, particularly by disrupting directed paths that connect regulatory nodes to their downstream targets. Redundant genes decreased the number of required driver nodes, and ordinary genes did not change the number of required driver nodes.

Type-II genes were classified based on their control requirements. Critical genes (zero in-degree, K_in_ = 0) were present in all driver node sets, redundant genes were absent from all driver node sets, and ordinary genes were present in some, but not all driver node sets.

We analyzed network connectivity using three measures: K (total degree), which represents total interactions and indicates network centrality; *K_in_* (in-degree), which shows incoming interactions that other genes could regulate; and *K_out_* (out-degree), which indicates outgoing interactions that influence different genes.

The CT score (*S*_CT_) incorporated these classifications with weighted importance. Type-I critical genes received a weight of 1 as they are essential for network stability. Type-II critical genes received a weight of 2 as they must always be controlled (K_in_ = 0). Redundant and ordinary genes received a weight of 0, reflecting their non-critical roles.

We calculated *S*_CT_ by multiplying each gene’s total degree (K) by its assigned weighted score (W):

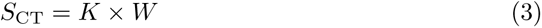

where *S*_CT_ represents each gene’s network impact score calculated from its degree and weight.

The final score was normalized using min-max scaling:

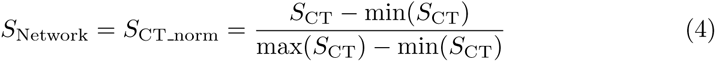

where min(*S*_CT_) and max(*S*_CT_) are the smallest and largest values across all genes, respectively.

#### 4.2.3 Analysis of Shared Genetic Basis Between Long COVID and Related Conditions

Disease-gene associations were compiled using five complementary databases: MalaCards [78], DISEASES [79], DISGENET [80], MedGen [81], and GenCC [82]. We systematically queried these databases for conditions associated with our identified genes, focusing on pathophysiological features that overlapped with Long COVID manifestations. Selection criteria included: (1) presence of immune/inflammatory components, (2) chronic/persistent symptoms, (3) multi-system involvement, and (4) metabolic or endocrine disruption. Conditions were categorized based on their primary pathophysiological mechanisms and potential relevance to Long COVID pathogenesis. Database selection was based on comprehensive coverage of rare and common conditions, including mechanistic annotations and regular curation of disease-gene relationships. The complete dataset of conditions and their database sources is provided in [Supplementary Data 4](https://github.com/SindyPin/Causal-Multiomics-Method/tree/main/SM/SM4).

#### 4.2.4 Enrichment Analysis (EA)

Our study conducted a comprehensive pathway EA on the risk, preventive, and network driver genes identified from our framework. The aim was to uncover the Biological Processes (BP), Cellular Components (CC), and Molecular Functions (MF) significantly associated with these genes. To ensure compatibility across various bioinformatics tools, we initially mapped gene IDs from Ensembl to Entrez ID using the org.Hs.eg.db database [74].

For the EA, we utilized well-established databases (GO [41], KEGG [42], and Reactome [83]). We prioritized enriched pathways based on statistical significance and relevance to established Long COVID literature. Pathways were considered significant when meeting all threshold criteria (p-value, p-adjust, and q-value *<* 0.05).

Furthermore, we examined the Long COVID context by conducting a meaningful literature review to identify potential symptoms that may be related to each enriched pathway, providing additional insights into the disease’s possible clinical implications.

We visualized the results using dot and network plots, representing enriched terms and molecular pathways clearly and intuitively.

#### 4.2.5 Gene Expression Clustering

We investigated Long COVID subtypes using gene expression data from our identified risk, preventive, and network driver genes. We determined the optimal number of clusters using the CancerSubtype package’s ConC algorithm [67], an unsupervised method for subtype discovery. The analysis used RNA-seq data detailed in Section 4.1.5. Moreover, we performed a grid search across hyperparameters, evaluating 2 to 5 clusters with a fixed seed of 5 for reproducibility.

After optimizing clustering parameters, we grouped Long COVID patients using the selected CC configuration. Cluster quality assessment involved calculating individual and group-wide silhouette widths. We selected the final number of clusters based on the highest ASW and balanced distribution of individuals across clusters. This clustering enabled the mapping of clinical data to analyze symptom prevalence within each subtype.

To assess cluster-specific symptom patterns, we conducted statistical significance testing. We applied Chi-square tests when expected cell counts in contingency tables exceeded 5. We used Fisher’s exact test for cells with lower expected counts, simulated p-values (workspace: 2e8) for symptoms, and simulated Chi-square tests for other clinical variables. Statistical significance was set at p-value *<* 0.05.

We then calculated symptom frequencies in absolute counts and relative percentages for each cluster, visualizing these distributions through comparative heatmaps.

More details about the entire framework can be found in ([Supplementary Data 9](https://github.com/SindyPin/Causal-Multiomics-Method/tree/main/SM/SM9))

## Data Availability

All data produced in the present study are available upon reasonable request to the authors

https://github.com/SindyPin/Causal-Multiomics-Method

## 5 Data Availability

Tissue-specific eQTL datasets were sourced from the https://gtexportal.org/home/datasets (Version 8, Ensembl 99, GRCh38). Long COVID GWAS datasets were obtained from Lammi et al., 2023 [73]. The WGS data, essential for the LD matrix calculation, was retrieved from GTEx (Ensembl 88, GRCh38), which can be obtained following the guidelines specified at https://gtexportal.org/home/protectedDataAccess. The Human Genes dataset for the causal relationship assessment, was obtained from the Ensembl Genes database (version 110, GRCh38), available at https://www.ensembl.org/. Long COVID RNA-seq data, used for critical gene discovery and molecular clustering, was derived from the Mount Sinai COVID-19 Biobank Study, available under GSE215865 at https://www.ncbi.nlm.nih.gov/geo/query/acc.cgi?acc=GSE215865. Finally, the PPI dataset, crucial for Long COVID network modeling, was retrieved from Vinayagam et al., 2011 [77].

## 6 Code Availability

The framework developed in this study to identify risk, preventive, and network driver genes is openly accessible for replication and further research at https://github.com/Causal-Multiomics-Method. This repository includes instructions for its use and a detailed guide to the data processing steps.

## Footnotes

1 Full link: https://sindypin.shinyapps.io/github/

